# Psychological distress and resilience in a sample of Adolescents and Young Adults with cancer during the COVID-19 pandemic

**DOI:** 10.1101/2021.03.29.21254529

**Authors:** Clare Jacobson, Rebecca Mulholland, Nicola Miller, Laura Baker, Daniel Glazer, Emily Betts, Louise Brown, Vera Elders, Olufunmilola Ogundiran, Robert Carr, Lee D Hudson

**Affiliations:** Guy’s and St Thomas’ NHS Foundation Trust, London; Sheffield Teaching Hospitals NHS Foundation Trust; Aberdeen and NHS Greater Glasgow and Clyde, Glasgow; University Hospitals Bristol NHS Foundation Trust; University College London Hospitals NHS Foundation Trust; Oxford University Hospitals NHS Foundation Trust; Royal Cornwall Hospitals NHS Trust; NHS Grampian; GOS UCL Institute and Child Health; GOS UCL Institute of Child Health

**Keywords:** Cancer, young adult, adolescent, psychological distress, mental health, resilience, COVID-19, SARS-CoV-2

## Abstract

**BACKGROUND:** Adolescents and young people (AYA) with cancer are at greater risk of psychological distress which can impact treatment. COVID-19 has resulted in changes to cancer care delivery and AYA have been disproportionately affected by economic and educational effects of COVID-19, potentially impacting on mental health. Understanding the impact of COVID-19 on AYA with cancer is important to inform care.

**METHODS:** Online survey of 16-24 year olds receiving cancer treatment at 8 cancer centres in the UK in December 2020. We measured: self-perceived increased anxiety since COVID-19, impact of COVID-19 on treatment, life and relationships and used the 8-item Patient Health Questionnaire(PHQ-8), 7-item Generalised Anxiety Disorder Scale(GAD) and the 2-item Connor-Davidson Resilience Scale(CD-RISC).

**RESULTS:** 112 AYA participated (17.8% of total eligible). 62.8% were female, 67.3% were 21-24 years. 83% were white. 59.8% had previously experienced mental health difficulties. 67.9% received cancer treatment during the pandemic and 33.9% were diagnosed during the pandemic. 78.6% reported COVID-19 having a significant impact on their life, 55.4% on their key relationship and 39.3% on their treatment. 79% reported experiencing some degree of increased anxiety since COVID-19.43.4% had moderate-severe PHQ-8 scores and 37.1% for GADS-7. Self-report of impact on life was associated with greater anxiety during COVID-19 and moderate-severe PHQ-8 score (OR 3.64, 95% CI 2.52 to 19.40, p <0.01; OR 5.23, 95%CI 1.65 to 16.56, p < 0.01). Impact on relationships was associated with greater anxiety and moderate-severe GADS-7and PHQ-8 score (OR 2.89, 95% CI 1.11 to 7.54, p = 0,03; OR 3.54, 95% CI 2.32 to 15.17, p<0.01; OR 2.42, 95% CI 1.11 to 5.25, p =0.03). Greater CD-RISC score was associated with lower risk of anxiety and mod-severe GADS-7and PHQ-8 scores (OR 0.58, 95%CI 0.41 to 0.81, p <0.01; OR 0.55 95% CI 0.4 to 0.72, p <0.01; OR 0.52, 95% CI 0.38 to 0.69, p <0.01)

**CONCLUSIONS:** We found high levels of psychological distress in AYA with cancer, which is important knowledge for clinical teams working with this age group. Perceived impact of COVID-19 on relationships and life was predictive of poorer mental health, with resilience a potential protective factor.

---

At the time of writing, the SARS-CoV-2 pandemic (COVID-19) has infected over 120 million people globally, and caused over 2.6 million deaths [1]. Though the highest risk factor for severe physical complications from SARS-CoV-2 infection in patients with cancer has been age, adolescents and young adults (AYA) with cancer have faced significant challenges as a result of the pandemic. As for many patients with cancer, care has been disrupted [2] and for some, initial presentations and diagnoses appear to have been delayed [3]. Concerns that AYA with cancer may be more at risk for severe complications of COVID-19, in particular those in treatment with immunocompromise [4], has led to some patients being advised to shield. Together with the broader population public health “lockdowns”, this has potentially led to isolation and loneliness [5] and household income and food insecurity [6]. Indeed, AYA patients with cancer form part of age group who have been disproportionately affected by economic and educational impacts of “lockdowns” with potentially significant knock-on effects on mental health [7].

Cancer is an important health burden for AYA globally [8], and around 2500 16-24 year olds diagnosed with cancer per year in the United Kingdom (UK) [9]. As a group, AYA with cancer are known to have high levels of psychological distress compared to both healthy peers and older adult cancer patients [10, 11] as well as patients of a similar age with other medical conditions.[12] Psychological distress in AYA cancer patients is associated with both increased morbidity and reduced concordance with treatment regimes which can adversely affect quality of life and potentially prognosis [13, 14].

How the additional pressures during the SARS-CoV-2 pandemic may have impacted on the psychological wellbeing of AYA with cancer is therefore an important question, and one for which there is currently limited published data. A recent multi-disciplinary position paper has called for timely research to understand the effect of COVID-19 on mental health as well as better understanding possible risk and protective factors [15]. This information is needed urgently to inform current AYA cancer patient needs and appropriate service delivery in the context of the pandemic. We studied psychological wellbeing in AYA cancer patients in the UK during COVID-19 using a longitudinal online survey, and present complete baseline data here.

## Methods

We recruited AYA patients who had received a cancer diagnosis between the ages of 16-24 years, and were currently engaged with cancer services at 8 UK centres to participate in an online study (Guy’ s and St Thomas’ NHS Foundation Trust, London, University College London Hospitals NHS Foundation Trust, London, Sheffield Teaching Hospitals NHS Foundation Trust, Oxford University Hospitals NHS Foundation Trust, University Hospitals Bristol NHS Foundation Trust, Royal Cornwall Hospitals NHS Trust, NHS Grampian, Aberdeen and NHS Greater Glasgow and Clyde, Glasgow). Participants needed to have undergone treatment within the last 2 years. We included some patients aged up to 30 years (but diagnosed between 16-24 years) to capture those who were still on long initial treatment protocols and those who had relapsed. We used the online survey platform onlinesurveys (Jisc) with the survey open for 2 weeks from 9-23^rd^ December 2020. Eligible patients were identified by local treatment teams and invited to take part in the survey via text message or email from a direct member of their NHS treating AYA team. The study is a longitudinal survey with 3 time points spanning 6 months, and currently only baseline data is available which we present here.

We asked participants to provide the demographics information on: current age group, age group at diagnosis, ethnicity, gender, eligibility in the past for free school meals (as a proxy for deprivation), living arrangements, diagnosis, treatment type and stage, whether they had been advised to shield, whether they were diagnosed or received active treatment during the pandemic, and whether they have a pre-existing mental health condition.

We used two validated self-report measures for psychological well-being (The 8-item patient health questionnaire (PHQ-8) and the 7-item Generalised Anxiety Disorder Scale (GADS-7)). The PHQ-8 has been established as a valid and reliable tool for assessing current depression in the general population and non-clinical settings [16, 17]. Further, studies have demonstrated a good internal consistency (crohnbach alpha = 0.82) in outpatient studies [17]. The eight items of the PHQ-8 each yield a score ranging from 0-3, providing a total severity score of 0-24. The cut-offs for a score of moderate distress is a total raw score of 10-14, moderately severe 15-19 and severe i20-24. The GADS-7 has also demonstrated high validity and reliability in the general population in assessing generalised anxiety disorder [18]. GADS-7 was also found as a reliable screening measure amongst cancer patients [19]. The seven items of the GADS-7 each yield a score ranging from 0-3, providing a total severity score of 0-21. Total raw scores of 5, 10, and 15 represent cut-off points for mild, moderate, and severe anxiety, respectively. We asked participants to report whether they had experienced more, less or no change in anxiety since COVID-19 began. We also asked participants to respond to a series of statements using a 6-point Likert scale (strongly disagree, disagree, neither agree nor disagree, agree, strongly agree). Statements were: 1) overall, COVID-19 has had a significant impact on my life. 2) COVID-19 has made having or having had cancer/a brain tumour harder than it otherwise would have been. 3) COVID-19 has made me feel anxious about returning to hospital for appointments or treatment. 4) COVID-19 has had an impact on my treatment and/or care. 5) COVID-19 has had a significant impact on my key relationships.

We also used the 2-item Connor-Davidson Resilience Scale (CD-RISC) as a brief self-rating questionnaire of resilience. The CD-RISC has demonstrated good reliability and validity in a number of populations, which include a sample of cancer patients and a non-clinical sample of teenage and young adult students [20]. A total raw score ranging from 0-8 is yielded with a higher score indicating greater resilience.

Analyses were conducted using STATA (version 16). We reported summary descriptive data as proportions or averages (reported as means with standard deviations (SD) for normally distributed data and median and IQR for non-normally distributed data). We generated binary variables for: 1) Likert agreement statements divided into any degree of agreement versus any degree of no agreement, 2) GADS-7and PHQ-8 moderate to severe versus mild or none; 3) a little or a lot more anxious since COVID-19 versus same or less. We then used logistic regression models (thus reporting odd’ s ratios (OR) with 95% confidence intervals) to test for associations between potential predictors of 1) GADS-7mod to severe, 2) PHQ-8 mod to severe and 3) more anxious since COVID-19. Multivariable logistic regression models were used to include variables found to have significant univariable associations.

The study received ethical approval by the London Central Health Research Authority and was approved within the Research and development departments at all eight NHS trusts.

## Results

We recruited 112 participants from 628 eligible patients across the 8 centres at the time of the survey (17.8%). Baseline participant characteristics are shown in table 1. The sample contained a greater proportion of females (n=71, 62.8%) and the most common age group was 21-24 years (n=76, 67.3%). 18 (15.9%) of participants had been eligible for free school meals. Ethnicity was 83% white, 8.9% black or Asian, 0.9% Chinese and 7.2% mixed or other. The main diagnostic groups were haematological, neurological, head and neck, and skin cancers (see table 1). 68.8% were advised to shield and 59.8% had previously experienced mental health difficulties requiring psychological input. 67.9% received cancer treatment during the pandemic and 33.9% were diagnosed during the pandemic. 78.6% reported COVID-19 having a significant impact on their life, 55.4% on their key relationship and 39.3% on their treatment. Figure 1 shows 79% of the sample (n=88) reported experiencing some degree of increased anxiety since COVID-19. 37.1% (n=42) scored in the moderate-severe range on the GADS-7 and 43.4% of the sample (n=48) scored in the same range on the PHQ-8. 38.0% (n=43) reported that having cancer/brain tumour had given them skills to cope better with COVID-19. 53.1% (n=59) reported that COVID-19 had made their experience of cancer/brain tumour harder. The median CD-RISC score was 6 (interquartile range 4-7).

**Table 1.**
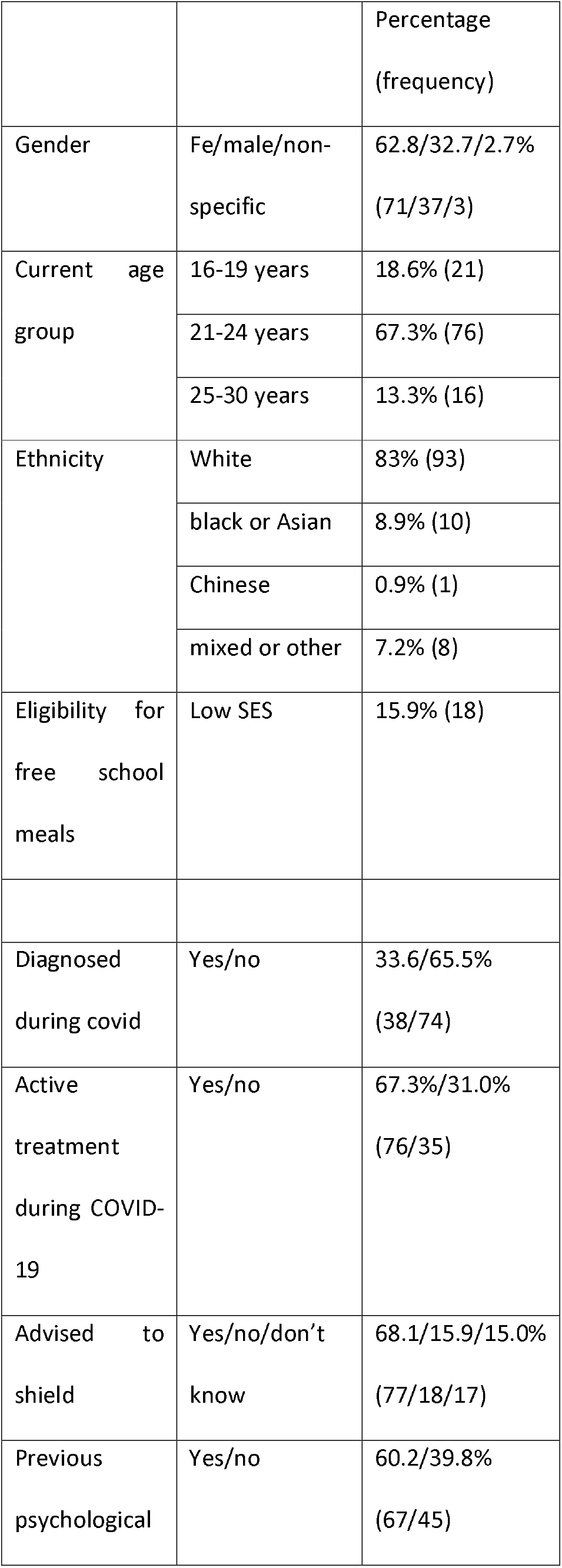
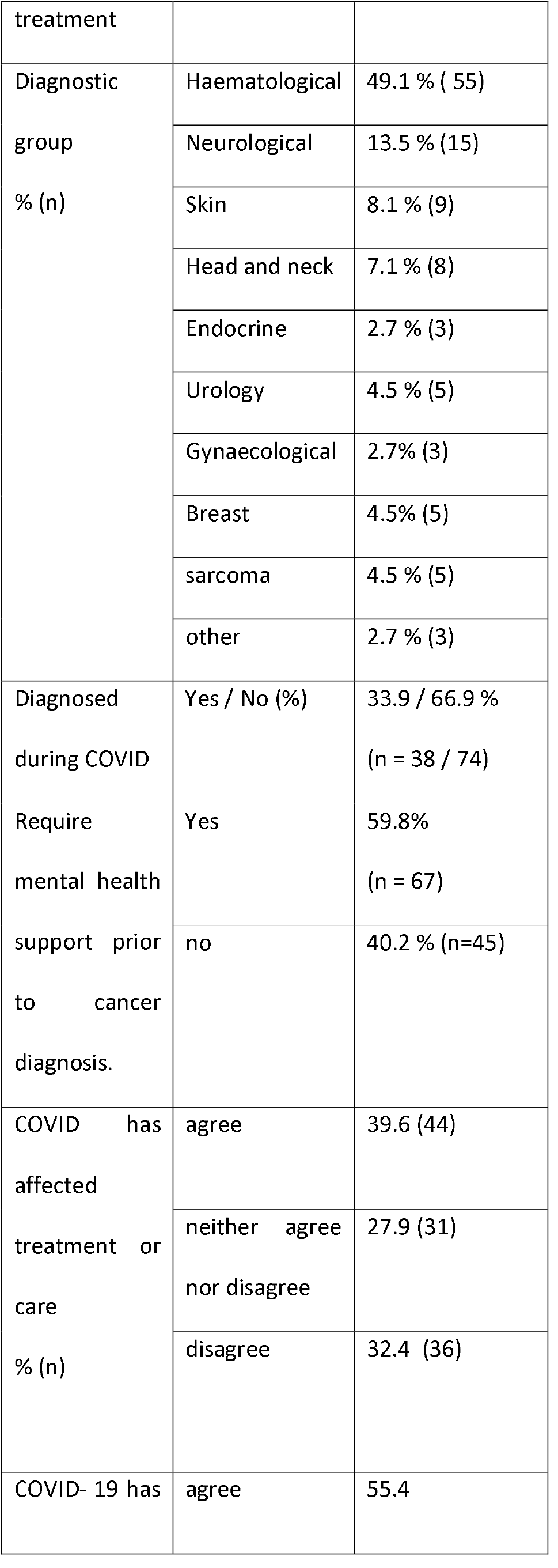
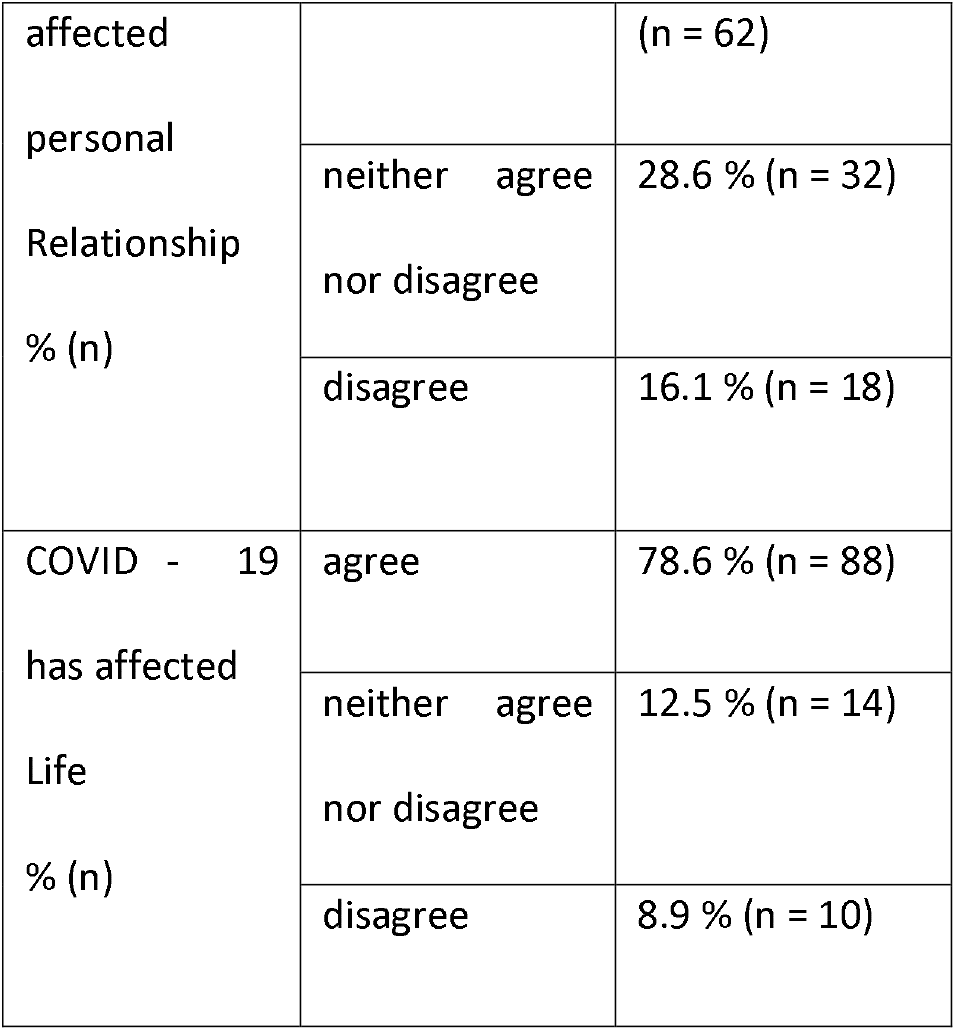

**Figure 1:**
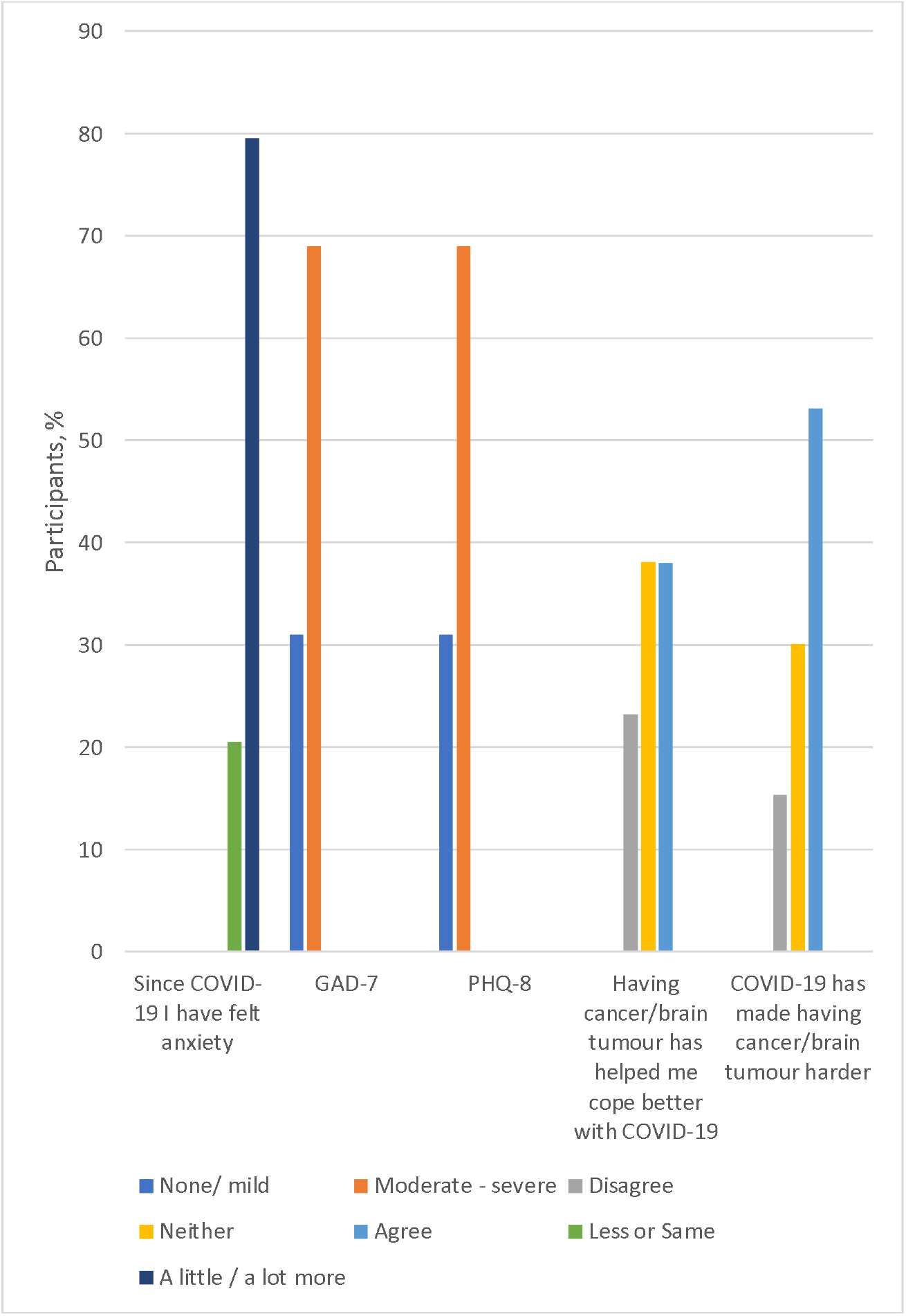
Graph showing proportions of participants for: A) degree of change in perceived anxiety since COVID-19; B) cut-off for severity of GAD-7; C) cut-off for severity of PHQ-8; D) agreement that cancer/brain tumour has helped coping with COVID-19; and E) agreement that COVID-19 has made having cancer/brain tumour harder.

Univariable logistic regression models from the sample for predicting A) more anxiety since COVID-19 B) mod-severe GADS-7 score and c) mod-severe PHQ-8 score are shown in table 2. No significant associations were found between age group, gender, ethnicity or free school meals and increased anxiety, mod-severe GADS-7 or PHQ. There were no significant associations found between the most common type of cancer (haematological) and increased anxiety, moderate-severe GADS-7 or PHQ. Presence of previous mental health difficulties was associated with greater odds both moderate-severe GADS-7 and PHQ-8 scores (OR 5.93, 95% CI 2.32 to 15.17, p <0.01; OR 5.92, 95% CI 2.46 to 14.26, p <0.01 respectively). Agreement that COVID-19 had impacted on life was associated with reporting being more anxious since COVID-19 and a moderate-severe PHQ-8 score (OR 3.64, 95% CI 2.52 to 19.40, p <0.01; OR 5.23, 95%CI 1.65 to 16.56, p < 0.01 respectively) but not moderate to severe GADS-7score. Agreement that COVID-19 had impacted on relationships was associated with reporting being more anxious since COVID-19 and a moderate-severe GADS-7and PHQ-8 score (OR 2.89, 95% CI 1.11 to 7.54, p = 0,03; OR 3.54, 95% CI 2.32 to 15.17, p<0.01; OR 2.42, 95% CI 1.11 to 5.25, p =0.03). The association between COVID-19 affecting life and moderate to severe PHQ-8 was robust when attenuated in a multivariable model with previous mental health difficulties (OR 5.31, 95% CI 1.58 to 17.92, p <0.01). Associations between COVID-19 affecting relationships with both moderate-severe GADS-7and PHQ-8 were robust when attenuated in a multivariable model with previous mental health difficulties (OR 3.81, 95% CI 1.55 to 9.36, p <0.01; OR 2.48, 95% CI 1.06 to 5.77, p = 0.04). There was a positive association between reporting that COVID-19 had made having or having had cancer harder with reporting being more anxious since COVID-19 and both moderate-severe GADS-7and PHQ-8 score (OR 20.3, 95% CI 4.45 to 92.43, p <0.01; OR 2.56, 95% CI 1.14 to 5.72, p 0.02; OR 2.5, 95% CI 1.14 to 5.45, p = 0.02 respectively). Greater CD-RISC score was associated with lower odds of the reporting of being more anxious than before COVID-19, and with lower odds of both mod-severe GADS-7and PHQ-8 scores (OR 0.58, 95%CI 0.41 to 0.81, p <0.01; OR 0.55 95% CI 0.4 to 0.72, p <0.01; OR 0.52, 95% CI 0.38 to 0.69, p <0.01)

**Table 2.** Univariable logistic regression of variables as predictors of A. little or more anxious than before COVID, B. A total GADS-7score indicating moderate or severe anxiety and C. A total GADS-7score indicating moderate or severe depression.

| A. A little or more anxious than before COVID |  |  |  |  |  |
| --- | --- | --- | --- | --- | --- |
|  |  | n | OR | 95% CI | p |
| Age group (vs. 16-19 years) | 20-24 years | 112 | 2.73 | 0.94 to 7.82 | 0.06 |
|  | 25-30 years |  | 8.61 | 0.94 to 78.67 | 0.06 |
| Gender |  | 111 | 2.02 | 0.77 to 5.33 | 0.16 |
| Ethnicity (vs. white) | Asian | 109 | 0.67 | 0.12 to 3.62 | 0.64 |
|  | Mixed |  | 0.11 | 0.01 to 1.31 | 0.64 |
| Free school |  | 112 | 1.88 | 0.39 to 9.05 | 0.43 |
| meals |  |  |  |  |  |
| Haematological malignancy |  | 111 | 0.97 | 0.38 to 2.49 | 0.96 |
| Active treatment during COVID |  | 111 | 1.94 | 0.75 to 5.00 | 0.17 |
| Diagnosis during COVID |  |  | 1.22 | 0.45 to 3.28 | 0.61 |
| Advised to shield |  | 112 | 2.46 | 0.78 to 7.75 | 0.12 |
| COVID-19 has made having or having had cancer harder |  | 111 | 20.3 | 4.45-92.43 | <0.01* |
| Agree or strongly agree COVID has impacted on life |  |  | 3.64 | 2.52 to 19.40 | <0.01 * |
| Agree or strongly agree COVID has impacted treatment |  | 111 | 2.15 | 0.78 to 5.98 | 0.14 |
| Agree or |  | 112 | 2.89 | 1.11 to 7.54 | 0.03 * |
| strongly agree<br>COVID has<br>impacted on<br>relationships |  |  |  |  |  |
| Previous<br>mental health<br>difficulties |  | 112 | 1.85 | 0.73 to 4.67 | 0.19 |
| Resilience |  | 112 | 0.58 | 0.41 to 0.81 | <0.01 * |
| B. GADS-7score indicative of moderate or severe anxiety |  |  |  |  |  |
|  |  | n | OR | 95% CI | p |
| Age group (v.s<br>16-19 years) | 20-24 years | 112 | 1.45 | 0.53 to 4.01 | 0.47 |
|  | 25-30 years |  | 0.5 | 0.11 to 2.37 | 0.38 |
| Gender |  | 111 | 1.63 | 0.70 to 3.81 | 0.26 |
| Ethnicity (vs<br>white) | Asian | 109 | 0.50 | 0.10 to 2.63 | 0.42 |
|  | Mixed |  | 0.76 | 0.07 to 8.65 | 0.82 |
| Free school<br>meals |  | 112 | 0.86 | 0.29 to 2.54 | 0.79 |
| Haematological<br>malignancy |  | 112 | 0.64 | 0.30 to 1.40 | 0.27 |
| Active<br>treatment<br>during COVID |  | 111 | 1.18 | 0.51 to 2.73 | 0.70 |
| Diagnosis<br>during COVID |  | 112 | 1.34 | 0.60 to 2.99 | 0.47 |
| Advised to |  | 112 | 2.11 | 0.63 to 7.04 | 0.22 |
| shield |  |  |  |  |  |
| COVID-19 has<br>made having or<br>having had<br>cancer harder |  | 111 | 2.56 | 1.13 to 5.74 | 0.02* |
| Agree or<br>strongly agree<br>COVID has<br>impacted on<br>life |  | 112 | 2.76 | 0.94 to 8.06 | 0.06 |
| Agree or<br>strongly agree<br>COVID has<br>impacted<br>treatment |  | 111 | 1.32 | 0.60 to 2.90 | 0.48 |
| Agree or<br>strongly agree<br>COVID has<br>impacted on<br>relationships |  | 112 | 3.54 | 1.54 to 8.16 | <0.01* |
| Previous<br>mental health<br>difficulties |  | 112 | 5.93 | 2.32 to 15.17 | <0.01 |
| Resilience |  | 112 | 0.55 | 0.4 to 0.72 | <0.01* |
| C. PHQ-8 score indicative of moderate or severe depression |  |  |  |  |  |

|  |  | n | OR | 95% CI | p |
| --- | --- | --- | --- | --- | --- |
| Age group (vs. 16-19 years) | 20-24 years | 112 | 1.89 | 0.69 to 5.22 | 0.22 |
|  | 25-30 years |  | 1.0 | 0.99 | 0.25 to 4.07 |
| Gender |  | 111 | 1.43 | 0.63 to 3.21 | 0.39 |
| Ethnicity (vs white) | Asian | 109 | 0.39 | 0.07 to 2.02 | 0.26 |
|  | Mixed |  | 0.58 | 0.05 to 6.64 | 0.66 |
| Free school meals |  | 112 | 1.47 | 0.53 to 4.10 | 0.46 |
| Haematological malignancy |  | 111 | 0.96 | 0.45 to 2.03 | 0.92 |
| Active treatment during COVID |  | 111 | 1.21 | 0.54 to 2.73 | 0.64 |
| Diagnosis during COVID |  | 112 | 1.24 | 0.56 to 2.75 | 0.58 |
| Advised to shield |  | 112 | 1.95 | 0.63 to 6.01 | 0.25 |
| COVID-19 has made having or having had cancer harder |  | 111 | 2.5 | 1.14 to 5.45 | 0.02* |
| Agree or strongly agree COVID has impacted on |  | 112 | 5.23 | 1.65 to 16.56 | <0.01* |
| life |  |  |  |  |  |
| Agree or<br>strongly agree<br>COVID has<br>impacted<br>treatment |  | 111 | 1.00 | 0.46 to 2.15 | 0.99 |
| Agree or<br>strongly agree<br>COVID has<br>impacted on<br>relationships |  | 112 | 2.42 | 1.11 to 5.25 | 0.03* |
| Previous<br>mental health<br>difficulties |  | 112 | 5.92 | 2.46 to 14.26 | <0.01 |
| Resilience |  | 112 | 0.52 | 0.38 to 0.69 | <0.01* |

## Discussion

In our sample of AYA cancer patients from 8 centres in the UK, we found high levels of reported psychological distress, with around 80% of respondents reporting some degree of increased anxiety since the beginning of COVID-19, and around 40% meeting moderate to severe cut-offs on both the PHQ-8 and GADS-7 respectively. This is concerning given the known impact that psychological distress can have on cancer treatment in AYA with cancer [13]. We believe this to be the first study to examine for participants’ own perception of COVID-19 impact on their treatment, lives and relationships. A majority of participants reported that COVID-19 had made a significant impact on their lives and their relationships, and had made their experience of cancer harder. Impact on life and relationships were found to be associated with psychological measures of depression and anxiety, in particular a reported impact on relationships associated with 3.5 odds of a moderate or severe GADS-7scores and 2.5 odds of a severe or moderate PHQ-8 score. Perhaps not surprisingly, previous mental health difficulties were associated with high PHQ-8 and GADS-7 scores in our sample. However interestingly in analyses, when previous mental health difficulties were adjusted for, the relationship between impact on relationships and GADS-7 and PHQ-8 remained robust. We were surprised that neither active treatment during COVID-19 nor diagnostic group were associated with increased distress, however we lacked data on treatment modality and this warrants further investigation. Importantly, we found reduced odds of both a report of being more anxious since COVID-19 and both moderate to severe GADS-7and PHQ-8 scores with greater resilience scores (CD-RISC) which represents a potentially important moderator to be considered by clinicians.

Our findings are similar to those reported by Kosir et al [21] who early in the pandemic similarly used an online survey to examine depression and anxiety using the PHQ-4 item scale for depression and a 2-item scale for anxiety in 177 AYA cancer patients aged 18-39 years in Slovenia and the UK during the early stages of the pandemic in April 2020. A higher proportion of our sample reported greater anxiety since COVID-19 and a greater proportion reported significant distress and/or anxiety. Differences could be explained by the timepoint within the pandemic (April 2020 compared to December 2020 for our study). Casanova et al [22] found increased distress in a paediatric and AYA sample was related to perceived risk of severe complications from COVID-19 for their health. Our sample did not find an association between perceived impact of COVID-19 on healthcare but it did for perceived impact of COVID-19 on general life and relationships. One potential mechanism for how relationships for AYA with cancer may have been affected is that during the pandemic, family or significant others were usually unable to accompany AYAs for treatment or visit, and young people may have been separated by lockdown and shielding. This may have caused distress, longer-term relational effects and reducing ‘connectedness’. Importantly, connectedness has been identified as a process that facilitates resilience [23], predicts young people’ s development of post-traumatic stress [24] and mediates psychological adjustment in young people with cancer [25]. The apparent protective effect of resilience for psychological distress in our sample is in keeping with the widely accepted concept that building patients’ resilience with a cancer diagnosis is crucial in mediating the psychological distress and coping throughout the cancer experience [26]. The findings in our sample suggests resilience may have a role in moderating the effects for CYP by clinical teams in the current context of the pandemic, and further enforces the need for a preventative models of psychology [15] for young people with cancer [25].

Our study has a number of strengths and limitations. We used validated psychological questionnaires, alongside pragmatic questions to identify specific perceived impacts during COVID-19. There were a greater proportion of females in our sample and age group predominance was disproportionate (with the majority being aged 22-24). However, we deliberately recruited up to the age of 30 because whilst we were interested in AYA patients (16-24) we did not want to exclude data from patients who were diagnosed in this age group but continued treatment for a longer period or had relapsed. The proportion of AYA who reported being eligible for free school meals is similar to nationally reported rates (13.6% in England[27]), suggesting representativeness of socio-economic status of the total population. We did not formally power our sample size, with the aim of recruiting as many AYA as possible. Though we recruited from 8 centres and targeted all eligible patients within those centres, our sample is small and less than 20% of eligible patients. This might have meant that our sample was not adequately powered to detect small effect sizes in univariable logistic regression models. The sample size also potentially leads to bias, in particular because AYA with higher levels of psychological difficulties may have been motivated to take part. Our sample is also cross-sectional and associations we have reported do not imply causation. Similarly, reported psychological measures may not have been temporally stable over time. That said, our study methodology is longitudinal and we will report at a future point on change over time.

## Conclusion

Within the constraints of our sample size, we believe that the findings in our survey provides important information for care of AYA with cancer since the pandemic began. Our data suggests that the impact of COVID-19 on relationships and overall life may be predictive factors for poorer mental health in AYA with cancer and an area to explore with patients to look for potential solutions and opportunities is to enhance resilience as a possible protective factor.

## Data Availability

The datasets analysed during the current study are available from the corresponding author on reasonable request.

## Declaration

The study received ethical approval by the London Central Health Research Authority (Committee’ s IRAS reference is 285244) and was approved within the Research and development departments at all eight NHS trusts. Participant consent to participate and to publish was sought. Funding for the online platform was provided by the Teenage and young adult cancer charity fund held by the Guy’ s and st Thomas’ NHS Foundation trust charity. The datasets analysed during the current study are available from the corresponding author on reasonable request.

## Author contributions

Clare Jacobson, Rebecca Mulholland, Nicola Miller and Laura Baker lead adolescent and young adult cancer psychology services in the NHS and conceived the idea for the study together. They all contributed to study design, completed the protocol and completed the ethical approval process. Clare Jacobson and Olufunmilola Ogundiran managed the data. Clare Jacobson and Lee Hudson conducted analyses and produced the first draft of the manuscript. All authors contributed to the final manuscript. Clare Jacobson and Rebecca Mulholland co-founded and co-chair the National Teenage and Young Adult Clinical Psychology Network for the UK. There are no conflicts of interest.

## Acknowledgements

We would like to acknowledge and thank the young people who took part in the survey and the staff at individual cancer centres who supported the survey and its communication to eligible participants. In particular, we would like to thank Gavin Maynard-Wyatt, Hannah Lind, Nicola Clapson, Amanda Copland and Louise Morris.

## List of abbreviations

AYA: adolescents and young adults
PHQ-8: Patient Health Questionnaire
GADS-7: 7-item Generalised Anxiety Disorder Scale
CDRS-2: 2-item Connor-Davidson Resilience Scale
COVID-19: SARS-CoV-2 pandemic

## Notes

### Competing Interest Statement

The authors have declared no competing interest.

### Funding Statement

Funding for the online platform was provided by the Teenage and young adult cancer charity fund held by the Guys and st Thomas NHS Foundation trust charity

### Author Declarations

The study received ethical approval by the London Central Health Research Authority and was approved within the Research and development departments at all eight NHS trusts. Participant consent to participate and to publish was sought.

### Summary of Updates

Authorship list submitted incorrectly first submission

